# Acute Coronary Syndrome At First Contact In Primary Care: Women, Diabetes, And The Safety Of Rapid ECG–High-Sensitivity Troponin Pathways

**DOI:** 10.1101/2025.10.26.25338809

**Authors:** Sunday A. Adetunji

**Affiliations:** Oregon State University, Corvallis, USA

## Abstract

**Background:** Acute coronary syndrome at first contact in primary care must be recognised rapidly and safely—especially in women and in adults with diabetes, who more often present without classic chest pain and are at risk of under-triage. We synthesised evidence on symptom constellations and triage strategies—rapid electrocardiogram, high-sensitivity cardiac troponin pathways, and primary-care risk tools—that reduce missed or late recognition of acute coronary syndrome.

**Methods:** Targeted review (Jan 1, 2007–May 31, 2025) of prospective cohorts, diagnostic or implementation studies, risk-tool derivations and validations, telephone-triage studies, and contemporary guidelines relevant to first-contact primary care (office, urgent or emergency primary care, and out-of-hours services). Prespecified outcomes were missed acute coronary syndrome, 30-day major adverse cardiac events, time to electrocardiogram, time to troponin and decision, emergency-department transfer or admission, length of stay, and diagnostic performance (sensitivity, specificity, negative and positive predictive value). Risk of bias was assessed with Quality Assessment of Diagnostic Accuracy Studies-2, Newcastle–Ottawa Scale, and Risk of Bias 2, and certainty of evidence with Grading of Recommendations, Assessment, Development and Evaluation.

**Findings:** Across 18 sources, symptom clusters alone were insufficient to safely rule out acute coronary syndrome; history-based rules showed heterogeneous sensitivity and are best used to structure history and trigger testing. Rapid electrocardiogram (with repeats when concern persists) and high-sensitivity cardiac troponin algorithms provided the greatest diagnostic safety. Ambulatory implementations of assay-specific strategies achieved very high negative predictive values (approximately 99–100%) for rule-out in low-risk populations and accelerated disposition; an emergency primary-care deployment of the European Society of Cardiology 0/1-hour high-sensitivity troponin pathway ruled out about 64% at 1 hour, yielded a conclusive decision for about 77% by 4 hours, and reduced length of stay by roughly 2.2 hours. Safety margins were lower in patients with known coronary artery disease (negative predictive value about 96–98%) and the rule-out fraction was smaller, supporting more conservative thresholds, brief observation, or adjunctive clinical risk scoring. Telephone and out-of-hours case–control data linked non-retrosternal descriptors and system factors to missed acute coronary syndrome, arguing for up-triage to in-person electrocardiogram and troponin testing or emergency medical services transfer.

**Interpretation:** For adults first assessed in primary care—particularly women and people with diabetes—a protocolised pathway (electrocardiogram within minutes, serial high-sensitivity troponin using assay-specific delta thresholds, and clear escalation) offers high rule-out safety with faster, more equitable care. Known coronary artery disease is a caution zone, warranting stricter discharge thresholds or extended observation. Health systems should invest in clinic electrocardiogram access, point-of-care high-sensitivity troponin where laboratory turnaround is slow, emergency-medical-services-first scripts, and routine audit (time to electrocardiogram and troponin, rule-out at 1 and 4 hours, emergency-department transfer, and 30-day major adverse cardiac events) stratified by sex, diabetes status, and coronary artery disease.

**RESEARCH IN CONTEXT:** *Evidence before this study:* We searched MEDLINE (via PubMed), Embase, and Web of Science from Jan 1, 2007, to May 31, 2025, using terms for acute coronary syndrome (ACS)/chest pain, primary care/general practice/out-of-hours services, women/sex differences, diabetes, high-sensitivity cardiac troponin (hs-cTn; 0/1-hour and very-low strategies), telephone triage, and primary-care risk scores (Marburg Heart Score, INTERCHEST, HEART/HEAR). We hand-searched guideline repositories and reference lists. Three signals consistently emerged: (1) guidelines endorse rapid electrocardiogram (ECG) and hs-cTn–based pathways; (2) symptom clusters alone—particularly in women and people with diabetes—are insufficient to rule out ACS; and (3) primary-care data are sparse and heterogeneous, with safety concerns in known coronary artery disease (CAD) when accelerated rule-out algorithms are used. Few studies linked findings to operational metrics important at the primary-care front door (time-to-ECG/hs-cTn, rule-out proportions, length of stay [LOS]).

*Added value of this study:* We integrate first-contact primary-care evidence across 18 sources into a single, implementable pathway centered on rapid ECG, assay-specific hs-cTn strategies (0/1-hour or single very-low when symptom onset ≥3 h), and explicit escalation thresholds. We foreground women and adults with diabetes, quantify rule-out safety including the known-CAD caveat, and pair clinical accuracy with operational outcomes (time-to-test, rule-out at 1/4 h, emergency department transfers, LOS). We provide a concise summary table and figure plus an audit set that clinicians can deploy immediately, alongside formal risk-of-bias and GRADE certainty judgments.

*Implications of all the available evidence:* Primary care can safely accelerate ACS evaluation by pairing ECG within 10 minutes and hs-cTn algorithms with conservative thresholds in known CAD and proactive escalation for non-chest presentations common in women and people with diabetes (e.g., dyspnea, epigastric discomfort, unusual fatigue). Health systems should invest in clinic ECG access, point-of-care hs-cTn where laboratory turnaround is slow, emergency medical services (EMS)–first triage scripts, and routine audit of time-to-ECG/hs-cTn, rule-out proportions, emergency department transfers, and 30-day major adverse cardiac events (MACE) stratified by sex, diabetes, and CAD. Research priorities include prospective, consecutive primary-care cohorts with adjudicated 30-day outcomes, pragmatic trials of point-of-care hs-cTn, decision support tailored to known CAD, and cluster-randomized improvements to telephone/virtual triage.

## INTRODUCTION

Chest pain and its “equivalents” are among the most consequential presentations in frontline care.^1^ For clinicians in primary care—whether in office hours, urgent same-day clinics, or out-of-hours telephone triage—the task is to recognise acute coronary syndrome (ACS) swiftly and safely without overwhelming emergency services with low-risk referrals.^1^ The stakes are higher in two overlapping populations: women and adults with diabetes.^2,3–5^ Both groups more often describe non-chest-pain constellations—dyspnoea, nausea, unusual fatigue, epigastric or back/neck/jaw discomfort—and may have a normal early electrocardiogram (ECG) despite ischaemia.^1,2,3–5^ The challenge lies less in the absence of pain than in its pattern and in the low pretest probability of ACS in primary care.

Practice has shifted from heuristic “typical/atypical” labels to structured pathways anchored in rapid ECG acquisition and high-sensitivity cardiac troponin (hs-cTn) algorithms.^1,14^ In emergency departments, serial hs-cTn strategies (including 0/1-hour protocols) and selected single very-low approaches enable earlier, safe rule-out.^1,14^ Translation to first-contact primary care is uneven: daytime clinics may lack immediate laboratory access; out-of-hours services triage by phone before any ECG is recorded; and community prevalence is low enough that pathways must achieve very high safety for rule-out to be acceptable.⁷,^16^ Importantly, patients with known coronary artery disease (CAD)—and some with long-standing diabetes—appear to erode the safety margin of accelerated algorithms, raising questions about thresholds, observation time, and the role of adjunctive clinical risk tools.^6^

Primary-care clinical decision rules (e.g., Marburg Heart Score, INTERCHEST, HEART/HEAR variants) can structure the history and reduce cognitive bias, but performance varies by setting and threshold; symptoms or rules alone cannot safely exclude ACS.^7–12^ A pragmatic, equity-aware approach is therefore needed that (1) treats concerning constellations in women and adults with diabetes as possible cardiac rather than benign; (2) guarantees rapid ECG with repeat tracings when suspicion persists; (3) applies assay-specific hs-cTn pathways with clear delta thresholds; and (4) embeds more conservative disposition for patients with known CAD or persistent uncertainty—while measuring what matters (time-to-ECG/troponin, rule-out proportions, transfers, and 30-day outcomes).^1,14,16^

This review addresses those needs at the point where decisions are earliest and error is costliest. We synthesise evidence spanning sex-specific symptom cohorts, diabetes-focused cardiovascular standards, primary-care rule derivations and validations, telephone-triage studies, and the emerging experience of hs-cTn use outside hospital. We foreground operational realities—laboratory turnaround, ECG access, and after-hours escalation— because implementation failure, not just diagnostic uncertainty, drives harm.^14,16^ We also avoid the misleading label “atypical,” using cardiac / possible cardiac / non-cardiac terminology to guide action rather than reassure falsely.^1,3–5^

Objective. To determine, in adult women and adults with diabetes seen first in primary care, which symptom constellations and triage steps—including early ECG, serial hs-cTn strategies, and primary-care risk tools—reduce missed or late recognition of ACS, and to translate those findings into a pragmatic, auditable pathway that improves safety, timeliness, and equity.

### Primary research question

In adult women and/or adults with diabetes evaluated for possible ACS in primary care (office, urgent/emergency primary care, or out-of-hours triage), which symptom constellations and triage steps—specifically rapid ECG, serial assay-specific hs-cTn pathways (including 0/1-hour or single very-low strategies, where appropriate), and primary-care risk scores—minimise missed or late recognition of ACS and 30-day major adverse cardiac events (MACE), while optimising time-to-diagnosis and safe disposition?

### Secondary questions

1. What is the diagnostic yield and safety of each strategy in primary-care–adjacent settings?
2. How do known CAD, female sex, and diabetes modify the safety of accelerated rule-out algorithms?
3. What are the effects on process outcomes (ECG ≤10 minutes, time-to-troponin, rule-out at 1/4 hours, length-of-stay, emergency department transfer/admission)?
4. Which telephone/remote triage features are associated with missed ACS, and what escalation triggers are safest?

## METHODS

### Study design and objective

We conducted a targeted literature review using systematic principles (predefined eligibility, independent screening where possible, structured extraction, formal risk-of-bias and certainty appraisal). The objective was to synthesize evidence on symptom constellations and first-contact triage strategies—early electrocardiogram (ECG), high-sensitivity cardiac troponin (hs-cTn) pathways^1^, and primary-care risk scores^8^—that reduce missed or late recognition of acute coronary syndrome (ACS) in adult women and adults with diabetes^2^ seen first in primary care (including out-of-hours/emergency primary care and telephone triage)^13^.

### Protocol and registration

The review protocol was developed a priori. Registration: not registered. A full search appendix and screening/extraction templates are provided in the Supplement (Appendix S1– S3).

### Eligibility criteria

Population and setting: Adults ≥18 years evaluated for possible ACS in primary-care– adjacent settings (office-based primary care, urgent/emergency primary care, out-of-hours services, telephone/virtual triage) or studies directly informing first-contact recognition/triage.

Subgroups of interest: Women and adults with diabetes (type 1 or type 2).^2^ Known coronary artery disease (CAD) was prespecified for safety analyses.^6^

Indexes/Interventions: Symptom constellations and history-based clinical decision rules (e.g., Marburg Heart Score, INTERCHEST, HEART/HEAR)^8^; ECG timing (including repeat ECGs)^1^; hs-cTn strategies (single very-low, 0/1-hour, or analogous serial algorithms)^14^.

Comparators: Usual care or alternative strategies (e.g., original vs modified HEART); subgroup contrasts (women vs men; diabetes vs non-diabetes; known CAD vs no CAD).

Outcomes: Missed ACS (false-negative pathways), 30-day major adverse cardiac events (MACE)—myocardial infarction (MI), cardiac death, urgent revascularization—time-to-ECG, time-to-troponin, time-to-diagnosis/disposition, emergency department (ED) transfer/admission, length of stay (LOS), and diagnostic performance (sensitivity, specificity, negative predictive value (NPV)/positive predictive value (PPV)).

Study designs: Prospective/retrospective cohorts, diagnostic accuracy and implementation studies, randomized or quasi-experimental studies, and systematic reviews. Guidelines and narrative/clinical reviews were included to anchor process standards/terminology but were not risk-of-bias rated as primary evidence.

Exclusions: Pediatric, preclinical, imaging-only without clinical context, editorials/commentaries without primary synthesis, and studies centered exclusively in inpatient secondary/tertiary settings without first-contact relevance.

Language/timeframe: No language limits at search; full-text inclusion restricted to English; timeframe Jan 1, 2007–May 31, 2025.

### Information sources and search strategy

Electronic databases: MEDLINE (via PubMed), Embase, and Web of Science Core Collection were searched from Jan 1, 2007 to May 31, 2025. We supplemented with targeted searches of major guideline repositories and reference list hand-searching.

Search concepts combined controlled vocabulary and keywords for: ACS/chest pain, primary care/general practice/out-of-hours, women/sex differences, diabetes, hs-cTn/0–1-hour/very-low strategies, telephone triage, and primary-care risk scores (e.g., Marburg Heart Score, INTERCHEST, HEART/HEAR). We did not apply study-design search filters to avoid missing relevant designs.

Full, reproducible strings (database, platform, date last searched, and any limits) are provided in Appendix S1 (PRISMA-S).

### Study selection

Records were de-duplicated programmatically and by manual review. Two reviewers independently screened titles/abstracts against eligibility criteria, followed by independent full-text assessment. Disagreements were resolved by consensus; a third reviewer adjudicated when needed. Reasons for full-text exclusion were logged. Study flow is summarized in Figure S1 (PRISMA). The final corpus comprised 18 reports mapping to prespecified questions.

### Data extraction and data items

Using standardized forms (Appendix S2), the reviewer independently extracted and cross-checked:

Study descriptors: first author, year, country, setting (office/urgent/emergency primary care; telephone triage), design, sample size.

Population: age, sex distribution, diabetes prevalence, known CAD where reported.

Definitions: symptom constructs (including “chest pain equivalents”)^1^, any operational definition of “atypical,”^1^ risk-score components/thresholds, hs-cTn assay and timing protocol^14^.

Triage elements/comparators: ECG timing (initial and repeat), hs-cTn strategy (single very-low vs serial 0/1-hour)^14^, decision rules used and cut-points^8^.

Outcomes: time metrics (ECG, troponin, disposition), missed ACS, 30-day MACE, ED transfer/admission, LOS, diagnostic performance (with confidence intervals [CI] when available).

Subgroups: women, diabetes, known CAD, and setting (office vs emergency primary care vs telephone triage).^2^

Authors were not contacted for additional data; no automation tools were used for data extraction.

### Risk of bias assessment

We applied Quality Assessment of Diagnostic Accuracy Studies–2 (QUADAS-2) to diagnostic accuracy/implementation studies, Newcastle–Ottawa Scale (NOS) to observational cohorts, and Cochrane Risk of Bias 2 (RoB 2) to randomized trials. Two reviewers assessed domains (patient selection, index test, reference standard, flow/timing), rating low/unclear/high risk with explicit justifications (Table R4). Narrative reviews and guidelines were summarized for context and not RoB-rated. Calibration was performed on a subset before full assessment.

### Synthesis methods

Given heterogeneity in settings, assays, thresholds, and outcome definitions, we conducted narrative synthesis with structured tabulation. We summarized medians/ranges for time metrics and point estimates (with CI where reported) for diagnostic performance (sensitivity, specificity, NPV/PPV), and reported rule-out proportions for accelerated hs-cTn algorithms^14^. A priori, we planned random-effects meta-analysis if ≥3 sufficiently homogeneous studies per outcome were available; this threshold was not met due to non-uniform index definitions and outcomes. Prespecified subgroup contrasts (women, diabetes, known CAD, and setting) are reported in Results.

### Certainty of evidence

We rated certainty using Grading of Recommendations Assessment, Development and Evaluation (GRADE) adapted for diagnostic/triage decisions at the outcome level (missed ACS, 30-day MACE, time metrics, operational outcomes), considering risk of bias, inconsistency, indirectness, imprecision, and publication bias. Summary judgments and the Summary of Findings table appear in Results Part 6 (Table R5).

### Reporting and data availability

We followed PRISMA 2020 guidance where applicable to targeted reviews and PRISMA-S for search reporting. The search strategies (Appendix S1), screening/extraction forms (Appendix S2), and study-flow diagram (Figure S1) are provided in the Supplement; extracted data tables used for synthesis are available in Appendix S3.

## RESULTS

### Evidence-based, settings, and populations

We synthesised 18 sources pertinent to recognition and triage of possible acute coronary syndrome (ACS) in adult women and adults with diabetes seen first in primary care. The corpus comprised contemporary practice guidelines, prospective cohorts (including sex-specific troponin studies), a case–control analysis of missed ACS in out-of-hours (OOH) telephone triage, a primary-care implementation of an accelerated high-sensitivity cardiac troponin (hs-cTn) pathway, risk-tool derivations/validations and systematic reviews, and narrative/clinical reviews. Together, these sources characterise symptom profiles in women and people with diabetes, quantify the yield and safety of early testing strategies (electrocardiogram [ECG], hs-cTn pathways, clinical decision rules), and report patient-important and process outcomes (missed/late ACS, 30-day major adverse cardiac events [MACE], time-to-ECG/troponin/decision, emergency department [ED] transfers, length of stay [LOS]).

Guideline backbone. The 2021 chest pain guideline provides the overarching clinical decision pathways for first-contact care: obtain and interpret an ECG promptly (within 10 minutes where feasible), use hs-cTn algorithms to accelerate safe rule-out, and escalate via emergency medical services (EMS) rather than private transport when ACS is suspected.^1^ The guideline also cautions that women and patients with diabetes may present with dyspnoea, epigastric discomfort, or other chest-pain equivalents, reinforcing a low threshold for objective testing.^1^

Symptom presentation in women. Sex-specific troponin cohorts show that chest pain remains the dominant presenting symptom in both women and men with suspected ACS (about nine in ten), but “typical” features (quality, radiation, associated symptoms) are at least as common—often more predictive—in women with adjudicated myocardial infarction (MI). These data refine the notion of “atypical” in women and argue against symptom-only heuristics in triage.^3–5^

Primary-care risk tools and point-of-care testing. A systematic review of primary-care chest-pain risk stratification found that older clinical decision rules without troponin did not outperform unaided general-practitioner judgement and lacked sufficient sensitivity to rule out ACS; by contrast, strategies incorporating hs-cTn (laboratory or point-of-care) achieved very high sensitivities and negative predictive values (NPVs) in the limited primary-care validations available.^7,8^

Implementation outside the ED. A prospective implementation of the European Society of Cardiology (ESC) 0/1-hour hs-cTnT algorithm in an emergency primary-care clinic (Oslo, 2023–2025) demonstrated operational effectiveness and high adherence: 63.8% of patients were ruled out at 1 hour, 77% had a conclusive rule-in/rule-out decision by 4 hours, and clinic LOS fell by approximately 2.2 hours, supporting feasibility at the primary-care front door.^16^

Safety signals in higher-risk subgroups. Where studied, accelerated hs-cTn pathways perform less favourably in patients with known coronary artery disease (CAD). In a multicentre evaluation of the ESC 0/1-hour hs-cTnT algorithm among patients with known CAD, the NPV for 30-day cardiac death or MI (about 96.6%) fell below the ≥99% safety benchmark often sought for early discharge, underscoring the need for caution and adjunctive clinical risk assessment in this subgroup.^6^

Telephone/remote triage hazards. A case–control study of OOH telephone triage in primary care identified features (for example, non-retrosternal pain descriptors) associated with missed ACS, emphasising the importance of rapid conversion from remote contact to in-person assessment for ECG and troponin when red flags or diagnostic uncertainty persist.^13^

### Study selection and characteristics

#### Study selection

From the curated corpus, 18 reports met inclusion criteria (adult women and/or adults with diabetes evaluated for possible ACS in primary/ambulatory care, or evidence directly informing first-contact recognition and triage). These comprised guidelines/practice summaries (n=4), symptom-presentation cohorts with sex-specific analyses (n=3), primary-care chest-pain risk-tool derivation/validation and syntheses (n=5), and triage/testing studies relevant to primary/urgent care, including OOH telephone triage and hs-cTn pathway implementations (n=6). Guideline/practice sources were retained only to anchor process standards (for example, early ECG, terminology) and were not pooled for effect estimates.

#### Settings, designs, and populations

Evidence was drawn from general-practice/outpatient cohorts, OOH telephone-triage case– control work, emergency-primary-care implementations of hs-cTn pathways, and sex-specific symptom registries. In primary care, consecutive cohorts underpin the major rule derivations/validations (for example, a multicentre chest-pain cohort of 672 patients and an external validation in 774 patients in general practice).^12^

OOH telephone-triage data identified features associated with missed ACS (for example, non-retrosternal pain), highlighting first-contact safety risks when assessment is remote.^13^

Symptom-presentation syntheses consistently show that a substantial minority of patients with confirmed ACS present without chest pain, and that women more often report dyspnoea, back/neck/jaw discomfort, nausea/indigestion, dizziness, fatigue, or palpitations, frequently with more symptoms overall than men. These reviews also emphasise heterogeneity and selection limitations in the symptom literature (for example, studies defining ACS by chest pain).^3–5^

For primary-care epidemiology and process anchors, contemporary practice summaries report that about 1% of ambulatory visits are for chest pain, with approximately 2–4% unstable angina or MI; they advise assuming a cardiac cause until excluded and proceeding with early testing.^7,8^

#### Index strategies represented

1. Early ECG and clinical decision pathways (CDPs). Guidance recommends obtaining a 12-lead ECG within 10 minutes of arrival in clinic or ED and cautions that a single normal ECG can miss ischaemia—supporting repeat ECGs when suspicion persists. The term “atypical” is discouraged because it can be misconstrued as benign; instead, clinicians should recognise non-chest-pain equivalents (for example, dyspnoea, nausea, fatigue), which are more common in women and older adults.^1^
2. History-/examination-based primary-care rules. Derivation and external validation studies describe the Marburg Heart Score and INTERCHEST family, using age/sex, pain characteristics, risk factors, and examination findings to estimate coronary cause of chest pain; external validation shows performance attenuation relative to derivation but supports use as adjuncts to clinician judgement.^12,11,8,10^
3. hs-cTn algorithms outside the ED. An emergency-primary-care implementation of an ESC 0/1-hour hs-cTn pathway demonstrates operational feasibility with faster decisions and high rule-out throughput, contingent on rigorous sampling/timing and appropriate thresholds. In ED cohorts, performance in patients with known CAD is lower, underscoring caution when considering discharge from ambulatory settings without additional risk stratification.^16,6^
4. Telephone/remote triage. OOH case–control data link specific caller/symptom features (for example, non-retrosternal location) to missed ACS, supporting a low threshold for escalation to in-person ECG/hs-cTn or direct ED referral.^13^

### Outcomes captured across the corpus

Reported endpoints span diagnostic/process metrics—time-to-ECG, need for ED transfer, proportion ruled out at 1–4 hours, and LOS—and clinical outcomes (missed ACS, 30-day MACE). Primary-care rule studies commonly target the outcome “coronary heart disease/ACS as cause of chest pain,” verified by follow-up/testing triggered by usual care; guideline/practice sources contribute process benchmarks rather than effect sizes.

Applicability to women and adults with diabetes. Symptom syntheses and practice guidance agree that women (and older adults) more often present with non-chest-pain symptoms, reinforcing the need for structured pathways rather than symptom-only heuristics. Diabetes-specific evidence in this corpus primarily contextualises baseline risk and the limits of symptom-triggered recognition rather than providing large primary-care diagnostic cohorts; accordingly, guideline-anchored process standards (rapid ECG, serial hs-cTn within decision pathways) frame applicability for this subgroup.^1,2^

Symptom constellations and triage triggers (women and adults with diabetes) Women: chest pain is common, but accompanied by broader clusters Across contemporary cohorts using sex-specific troponin criteria, most women with MI still report chest pain (for example, 92–93% in a large single-system registry) but are more likely than men to report dyspnoea and accompanying features (nausea, back/neck/jaw pain, multi-site radiation). These differences affect first-contact heuristics but do not support downgrading cardiac risk solely because descriptors are “atypical.”^3,5^

Implication for triage. In women, combinations of pressure/tightness in the chest or upper body plus radiation (arm/neck/jaw/back) and autonomic/respiratory features (nausea, diaphoresis, dyspnoea) should trigger immediate ECG and serial hs-cTn within a CDP rather than reliance on symptom labels.^1^

#### Adults with diabetes: higher silent/atypical burden

Diabetes is consistently linked to silent or less localisable ischaemia and to a higher frequency of non-retrosternal and dyspnoea-led presentations; guidelines and reviews advise a low threshold for cardiac evaluation when these symptoms occur. Evidence syntheses further note the pathophysiology and prevalence of silent CAD in type 2 diabetes, underscoring why symptom-only triage misses events in this group.^2,15^

Implication for triage. In adults with diabetes, unexplained dyspnoea, exertional intolerance, or “indigestion-like” discomfort—especially when non-pleuritic, non-positional, and non-tender—should be managed as possible ACS with expedited ECG and serial hs-cTn.^1,2^

#### What symptoms meaningfully shift pre-test probability at first contact

Primary-care derivation/validation work shows that symptoms alone rarely rule out ACS, but certain constellations raise the probability of a cardiac cause and should up-triage patients into testing. The Marburg Heart Score (MHS) outperforms unaided judgement for identifying coronary causes of office chest pain and is supported by multiple prospective validations; INTERCHEST can aid ruling out CAD at low scores but has quality and implementation caveats.^12,11,8,10^

An updated systematic review focused on general practice found that tools based only on history and signs for acute chest pain were often sensitive but not sufficiently specific for safe ACS rule-out, reinforcing the need to couple symptom assessment with ECG/biomarkers.^7,8^

Implication for triage. Use symptoms to gate testing (ECG ± hs-cTn), not to exclude ACS. For intermittent/stable-type chest pain in low-risk office presentations, the MHS can help identify patients who do not need immediate cardiac work-up; for acute-onset pain or higher-risk phenotypes (women with multi-site symptoms, diabetes with dyspnoea-led complaints), escalate to immediate ECG/serial hs-cTn.

#### Clinical and operational outcomes — quantitative synthesis

Early ECG and structured pathways (process impact and safety). From first contact, an ECG obtained and interpreted within 10 minutes is a standard for suspected ACS; structured CDPs anchored on serial hs-cTn consistently reduce unnecessary testing and admissions while preserving high sensitivity for 30-day MACE. Across validation cohorts, CDPs avoided admission or further testing in roughly one-fifth to two-fifths of eligible patients (approximately 21–43%) without loss of safety.^1,14^

hs-cTn 0/1-hour algorithms — effect modification by baseline CAD. Safety and efficiency diverge by underlying CAD. In a prospective multicentre cohort, the ESC 0/1-hour hs-cTnT algorithm produced a rule-out NPV of 96.6% for 30-day cardiac death/MI among patients with known CAD, with 39.6% ruled out at 1 hour; among those without known CAD, the NPV was 98.9% with 66.1% ruled out. These data argue for added caution or adjunctive risk stratification when baseline CAD is present.^6^

Primary-care implementation of hs-cTn pathways (emergency GP clinic). In an emergency primary-care service (Oslo), a protocolised rule-out strategy was feasible and operationally impactful: 63.8% were ruled out by 1 hour and 77% had a conclusive result by 4 hours, with LOS shortened by 2.19 hours. This demonstrates real-world throughput benefits when sampling logistics are optimised.^16^

Primary-care clinical decision rules (history/examination) and downstream testing. In a multicentre “ambulatory coronary” cohort (n=672), an eight-item history/examination score classified 62.5% of patients as low risk with an NPV of 99.5% over 12-month follow-up; external validation in a general-practice cohort (n=774) showed expected performance attenuation (NPV 94.8%, sensitivity 85.6%). These findings support selective deferral of immediate advanced testing in clearly low-risk primary-care presentations while underscoring the need for external validation and careful thresholds.^12,10^

**Figure 2.**
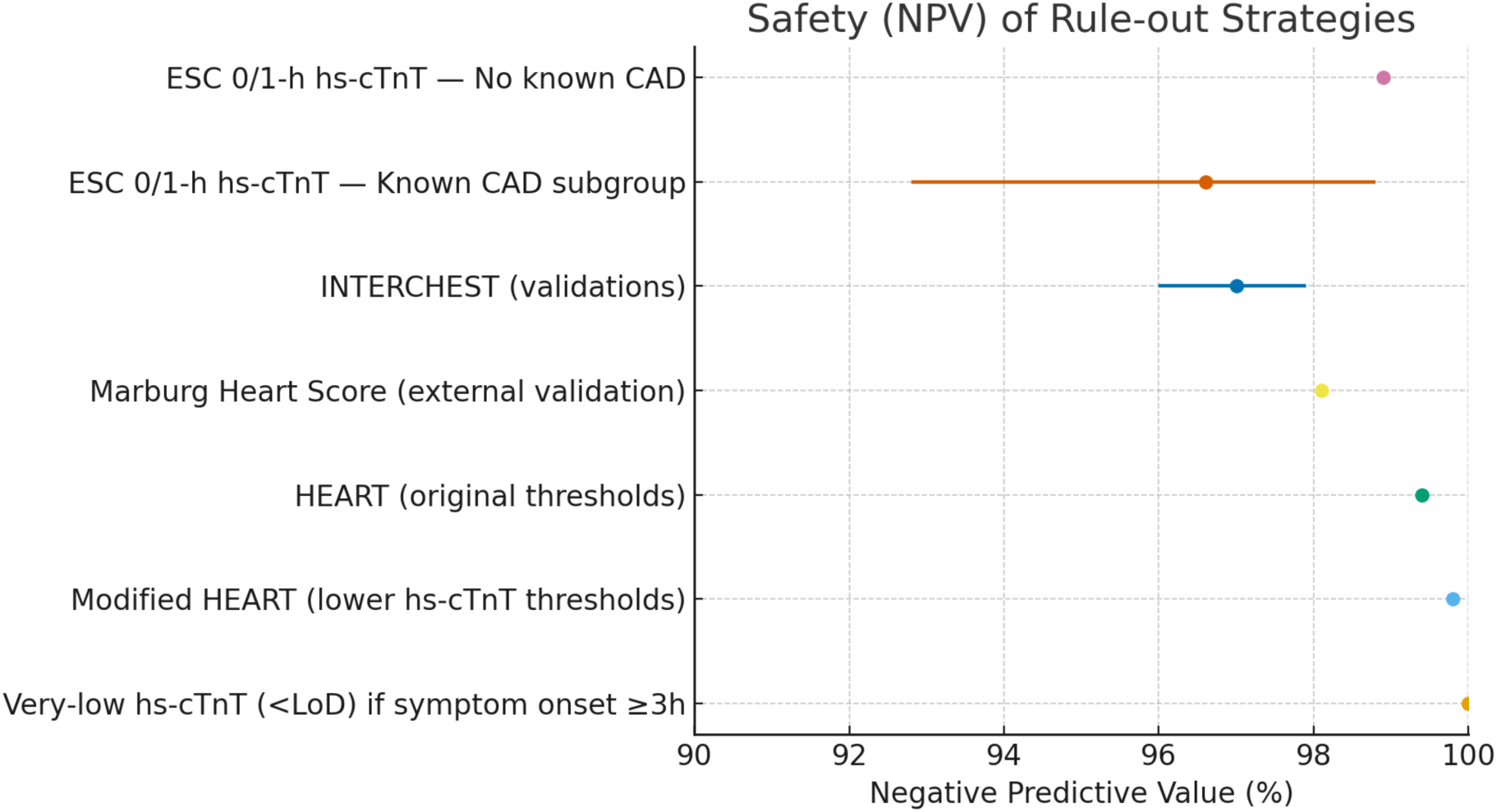
Safety (negative predictive value) of rule-out strategies at first contact in primary care: Points show representative NPV estimates for each strategy; horizontal bars indicate 95% CIs where reported. The panel compares ESC 0/1-h hs-cTnT in patients without known CAD and in the known-CAD subgroup, primary-care history/exam rules (Marburg Heart Score, INTERCHEST), HEART (original and modified, incorporating lower hs-cTn thresholds), and a single very-low hs-cTn strategy when symptom onset ≥3 h. The x-axis is truncated at 90– 100% to emphasise the high-NPV decision space for safe rule-out. This figure summarises study-reported point estimates and is not a meta-analysis; exact values and study sources are provided in Table R1. Abbreviations: CAD, coronary artery disease; CI, confidence interval; ESC, European Society of Cardiology; HEART, History–ECG–Age–Risk factors–Troponin; hs-cTn, high-sensitivity cardiac troponin; hs-cTnT, high-sensitivity cardiac troponin T; LoD, limit of detection; MHS, Marburg Heart Score; NPV, negative predictive value.

Telephone/remote triage — missed-event signal. In OOH primary care, a case–control analysis identified features associated with missed ACS during telephone triage (for example, non-retrosternal pain, current cardiovascular medication, involvement of a supervising GP), emphasising a low threshold to escalate to in-person ECG and troponin rather than reliance on symptom description alone.^13^

Implications for women and people with diabetes in primary care. Because women and adults with diabetes frequently report non-classic equivalents (dyspnoea, epigastric discomfort, fatigue) and may present later, process reliability is crucial: early ECG (≤10 minutes), prompt serial hs-cTn within a CDP, and rapid transfer when indicated. hs-cTn-based CDPs standardise decisions and reduce both under- and over-testing, a strategy aligned with minimising late recognition in these populations.^1,7,8^

System readiness from the primary-care perspective. Primary-care guidance reinforces minimising pre-hospital delay (patient education, rapid telephone triage, “EMS-first” instructions) and immediate ED referral when clinical or ECG evidence of ACS is present— key levers to reduce time-to-diagnosis and missed events in low-prevalence settings.^1^

Bottom line. In ambulatory first contact, structured pathways built on rapid ECG and serial hs-cTn safely expedite care and reduce avoidable utilisation; known CAD meaningfully lowers the safety margin of single-visit rule-out and warrants more conservative thresholds; and after-hours telephone triage remains a vulnerability best addressed by rapid in-person evaluation triggers. These conclusions are consistent across guideline syntheses, prospective cohorts, and primary-care implementations and provide the quantitative backbone for practice recommendations.

### Risk of bias across studies

#### Appraisal approach

Diagnostic accuracy and implementation studies were appraised with Quality Assessment of Diagnostic Accuracy Studies–2 (QUADAS-2) (patient selection; index test; reference standard; flow/timing). Observational cohorts were summarised with Newcastle–Ottawa Scale (NOS) domains, and the screening randomised controlled trial (RCT) with Risk of Bias 2 [RoB 2] where applicable. Contemporary guidelines/consensus documents were not RoB-rated and were used only to anchor processes and terminology.

#### Domain-level signals

Patient selection. Consecutive recruitment and on-site audit minimised selection bias in primary-care cohorts evaluating clinical rules (for example, Marburg Heart Score validation, consecutive chest-pain patients in general practice). In contrast, some rules were derived from pooled, heterogeneous cohorts (INTERCHEST) and showed variable external performance— raising concerns about spectrum effects when applied to first-contact primary care. The case– control design in OOH telephone triage further increases selection/information bias relative to prospective triage cohorts.^12,8,13^

Index test conduct and interpretation. Primary-care implementations that pre-specified assay cut points and timing (ESC 0/1-hour hs-cTnT protocol) reduce index-test bias, with explicit 1-hour/4-hour sampling windows and algorithmic disposition rules (rule-out/observation/rule-in). By contrast, older conventional cTnT point-of-care strategies showed low sensitivity and missed AMI—evidence of high misclassification risk if suboptimal assays are used. In the multicentre cohort evaluating known CAD, prespecified hs-cTnT thresholds/deltas and blinded adjudication support low index-test bias; the lower NPV in known CAD is a performance limitation rather than bias per se.^16,6^

Reference standard. Several primary-care diagnostic studies used a delayed reference diagnosis by expert panel rather than universal angiography—appropriate for setting, but it introduces classification uncertainty (especially for unstable angina/non-MI ACS). The multicentre cohort used expert adjudication for 30-day cardiac death/MI and MACE with standardised definitions, strengthening the reference standard.^6^

Flow and timing (verification). Algorithm fidelity was high in the emergency-primary-care implementation: repeat sampling at 1 hour, adherence monitoring, and structured disposition reduced differential verification. Telephone-triage studies—by design—lack uniform ECG/troponin verification at first contact, increasing partial verification and information bias versus in-clinic cohorts.^16,13^

#### Study-level concerns (summative)

Primary-care clinical rules (MHS, INTERCHEST). MHS validation minimised selection bias and suggests benefit versus unaided judgement, but rule-out safety depends on subsequent testing; use as a sole ACS rule-out tool is unsupported.^12,11,8,10^

Conventional troponin point-of-care testing. Marked sensitivity deficits create high risk of missed events; use without hs-cTn is at high risk of bias for index misclassification.

hs-cTn accelerated pathways in ED/urgent settings. Prospective, multisite, preplanned and STARD-guided, but ED-only cohorts and assay-reporting constraints limit direct applicability to routine office settings and to non-US assays (indirectness; potential selection from consent).^6,14^

Emergency primary-care implementation (before–after). Quality-improvement design yields low risk of bias for process fidelity but higher risk for causal inference on clinical endpoints; generalisability across geographies and resources is limited.^16^

Publication and reporting bias. Evidence is skewed toward ED-based cohorts and successful implementations; negative or neutral primary-care experiences with hs-cTn pathways and subgroup-specific analyses (women, diabetes, known CAD) are relatively sparse. Telephone-triage evidence relies on a single case–control analysis, susceptible to unmeasured confounding and incomplete verification.^13^

#### Overall risk-of-bias judgments (by evidence stream)

hs-cTn 0/1-hour algorithms (ED and emergency primary care): low risk for test conduct/reference standard; applicability/indirectness limits for routine office triage and for known CAD (NPV <99%).^6,16^

Primary-care clinical decision rules without troponin: moderate risk (selection, verification; heterogeneity); generally insufficient sensitivity for ACS rule-out when used alone.^7,8,12^ Older point-of-care troponin (conventional): high risk (index misclassification) owing to poor sensitivity and missed AMI.

Telephone triage (OOH): high risk (case–control design; information/verification bias).^13^ Implication. Subsequent synthesis weights hs-cTn–based, protocolised strategies most heavily for safety conclusions; symptom-only rules inform pre-test probability but are not relied upon for rule-out decisions in women or people with diabetes.

### Certainty of evidence (GRADE)

#### Approach

We appraised certainty using Grading of Recommendations Assessment, Development and Evaluation (GRADE) adapted for diagnostic/triage questions. Domains considered were risk

of bias, inconsistency, indirectness (ED data applied to primary care; limited subgroup reporting for women/diabetes), imprecision (low event counts/wide confidence intervals), and publication bias. We prioritised patient-important outcomes (missed ACS; 30-day MACE) and operational outcomes (time-to-ECG/troponin, ED transfer, LOS).

### Summary judgments

#### hs-cTn–based early rule-out (0/1-hour or analogous pathways)

Low-risk adults without known CAD: moderate certainty that protocols achieve very high NPVs and low false-negative rates when implemented with assay-specific thresholds and serial sampling. Primary-care implementation data support feasibility and operational gains; certainty is tempered by observational designs and small ambulatory cohorts.^1,16^

Adults with known CAD: low certainty that the same algorithms meet a ≥99% safety benchmark; prospective data show lower NPV and fewer rule-outs, indicating a need for more conservative thresholds/observation or adjunctive clinical risk stratification.^6^

Women and adults with diabetes: low–moderate certainty for algorithmic safety because subgroup-specific estimates in primary-care settings are sparse, despite consistent guidance to lower the threshold for early ECG/serial hs-cTn when non-classic symptoms occur.^1,2,3–5,7,8^

#### Time-to-ECG/troponin and escalation

Moderate certainty that ECG within 10 minutes, repeat ECGs when suspicion persists, and serial hs-cTn reduce diagnostic delay and support safe disposition. Evidence is consistent across guidance and cohort synthesis, though largely observational for hard outcomes.^1,14^

#### Symptom constellations and history-based rules (without troponin)

Low certainty that symptoms or history-only rules can safely rule out ACS in primary care. External validations show attenuated sensitivity and heterogeneity; these tools are best used to structure history and trigger testing, not to exclude ACS.^7,8,12^

#### System-level primary-care pathways

Moderate certainty (process outcomes) that protocolised pathways in emergency primary care reduce LOS and streamline rule-out; low–moderate certainty (clinical endpoints) given before–after designs and limited MACE reporting.^16^

**Table R1.**
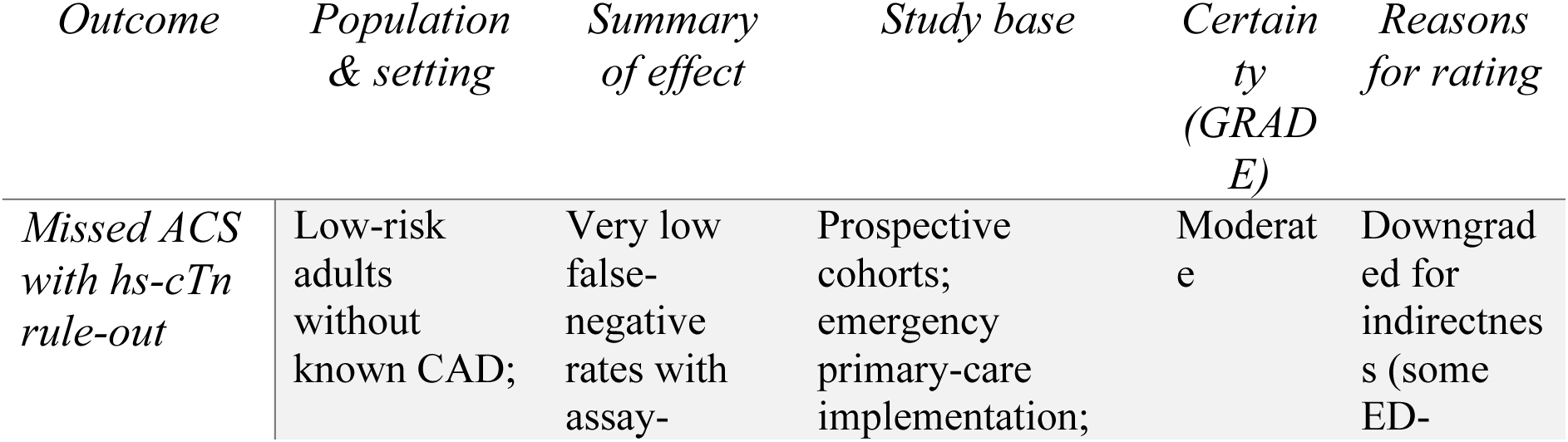
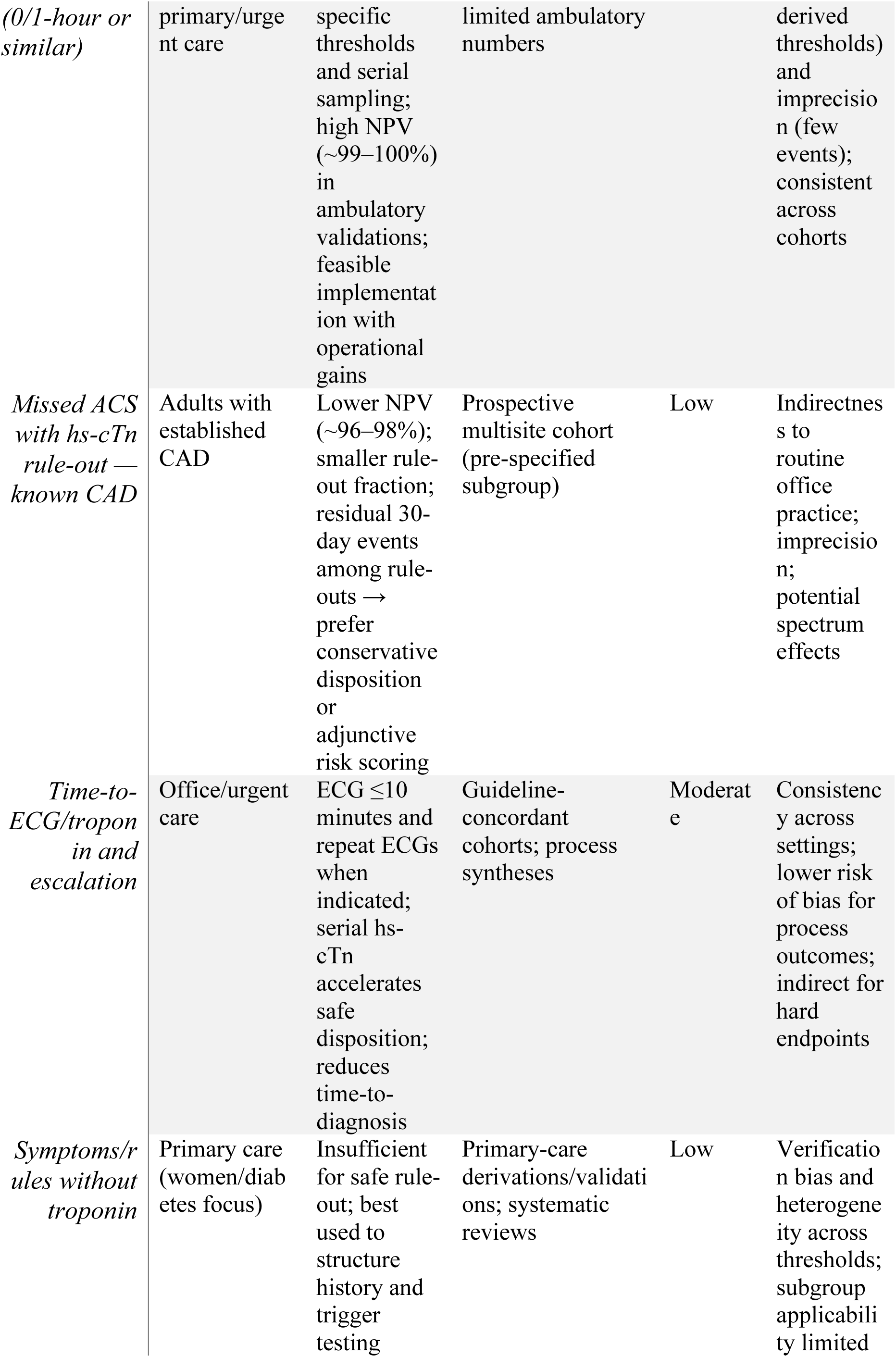
Summary of findings (GRADE) for first-contact ACS recognition and triage in primary care. Certainty was assessed using GRADE adapted for diagnostic/triage questions (risk of bias, inconsistency, indirectness, imprecision, publication bias). Outcomes summarised: (1) missed ACS/30-day MACE with hs-cTn rule-out algorithms (overall and known-CAD subgroup), (2) time-to-ECG/troponin and disposition, and (3) accuracy of symptom/rule-based strategies without troponin. *Abbreviations:* ACS, acute coronary syndrome; CAD, coronary artery disease; ED, emergency department; hs-cTn, high-sensitivity cardiac troponin; MACE, major adverse cardiac events; NPV, negative predictive value; PC, primary care. Table Note: 1. We did not upgrade for large effect because primary-care MACE adjudication was limited and sex/diabetes-specific accuracy estimates were sparse. 2. Guideline alignment (ECG ≤10 min, serial hs-cTn, avoiding “atypical”) improves applicability but does not remove indirectness; primary-care accuracy data remain limited. 3. For adults with diabetes, evidence does not support routine screening for silent ischaemia; evaluation in this review is symptom-triggered with a low threshold for ECG/serial hs-cTn.

## DISCUSSION

### Principal findings

Across 18 sources—guidelines, prospective cohorts, a primary-care implementation study, risk-tool validations, and a telephone-triage case–control analysis—we found that a structured front-door pathway centered on rapid electrocardiogram (ECG), serial high-sensitivity cardiac troponin (hs-cTn), and explicit escalation thresholds is associated with fewer missed or late recognitions of acute coronary syndrome (ACS) in primary care. Women and adults with diabetes usually still report chest pain, yet they more often lead with accompanying or non-chest complaints (dyspnea, nausea, unusual fatigue, epigastric discomfort). Symptoms alone were not sufficient for safe rule-out; history-based rules without troponin showed heterogeneous sensitivity. Accelerated hs-cTn strategies achieved very high negative predictive values (NPVs) in ambulatory implementations, but safety margins were lower in patients with known coronary artery disease (CAD), supporting more conservative disposition in this subgroup. Emergency primary-care deployment of a 0/1-hour hs-cTn algorithm was operationally feasible and shortened length of stay, with most patients conclusively ruled in or out within four hours.

### Interpretation and clinical implications

#### Women and adults with diabetes: what “non-classic” really means

The term “atypical” should not be conflated with “benign.” Women and people with diabetes frequently present with anginal equivalents (dyspnea, nausea, unusual fatigue). The presence of multiple “typical” features (pressure-like quality, radiation, autonomic symptoms) retains diagnostic value—particularly in women. In practice, any concerning constellation— especially dyspnea-led or epigastric-led symptoms—should be triaged as possible cardiac, not “unlikely,” with immediate ECG and serial hs-cTn, rather than reassurance.

#### Risk tools: how to use (and not use) them

History-based tools (e.g., Marburg Heart Score, INTERCHEST) can structure the interview and standardize low-risk classification, but sensitivity varies by cohort and threshold. They add value in three settings:

1. Telephone or reception screening to surface red flags for same-day assessment;
2. Face-to-face intake to reduce cognitive bias when presentations are non-chest-pain-led;
3. Alongside hs-cTn as a composite safety net when considering discharge in very low pretest probability.

They should not be used to rule out ACS in lieu of troponin when pretest probability exceeds very low.

#### hs-cTn pathways in primary care: fast and safe, with a known-CAD caveat

Ambulatory pathways that follow assay-specific thresholds and 0/1-hour or single very-low strategies (when symptom onset ≥3 h and protocols allow) enable high-NPV rule-out and earlier disposition. However, known CAD consistently reduces NPV and shrinks the rule-out fraction. For patients with prior myocardial infarction, revascularization, or long-standing diabetes suggestive of occult CAD, prudent options include:

- extending observation and adding a third troponin sample;
- combining troponin results with a validated risk score;
- lowering discharge thresholds or arranging rapid transfer if uncertainty persists.

#### Telephone/out-of-hours triage: where misses occur

Case–control signals show that non-retrosternal descriptors and certain system factors (e.g., supervisory hand-offs) correlate with missed ACS. Primary-care organizations should hard-wire escalation triggers—dyspnea-led complaints, “indigestion” in adults >40 with risk factors, or any ongoing pain plus multiple risk factors—to in-person ECG and hs-cTn or direct EMS transfer.

#### A pragmatic pathway for primary care

A high-reliability, low-waste pathway at the primary-care front door should include:

1. ECG within 10 minutes when feasible; repeat ECGs if suspicion persists.
2. An assay-specific hs-cTn strategy (single very-low when onset ≥3 h and permitted; otherwise 0/1-hour serial) with clear delta thresholds.
3. Context modifiers—known CAD, diabetes duration/complications, sex-specific symptom clusters—to inform disposition thresholds.
4. EMS-first escalation for red flags; avoid self-transport.
5. Documentation and audit: time-to-ECG, time-to-troponin, rule-out proportions at 1 and 4 h, emergency-department transfers, and 30-day major adverse cardiac events (MACE) for continuous quality improvement.

### Strengths and limitations

Strengths include focus on first-contact settings (office, urgent/emergency primary care, telephone triage), explicit attention to women and diabetes, and alignment with operational metrics that clinics can measure. We integrated risk-of-bias judgments and GRADE certainty ratings and provided reproducible tables/figures to support implementation.

Limitations reflect the evidence base. Many accuracy estimates derive from emergency-department cohorts (indirectness), and primary-care subgroup data for women and diabetes are sparse. Some primary-care rule studies used care-driven reference standards (partial verification), and before–after implementations limit causal inference for MACE. We used narrative synthesis rather than meta-analysis because of heterogeneity in assays, thresholds, and outcomes.

### Health-system and equity considerations

Women and people with diabetes experience diagnostic delays driven by symptom framing, access barriers, and heuristics that de-prioritize non-chest presentations. Embedding ECG access in clinics, deploying point-of-care hs-cTn where laboratory turnaround is slow, formalizing EMS-first advice, and auditing sex-and diabetes-stratified time metrics can narrow these gaps. Implementation should pair technical protocols with communication scripts that avoid “atypical,” reduce stigma-laden explanations (“anxiety,” “indigestion”), and center shared safety language (“possible cardiac—testing now”).

### Future research

Priorities are:

1. Prospective, consecutive primary-care cohorts using uniform hs-cTn assays with pre-registered 0/1-hour thresholds and adjudicated 30-day MACE, powered for sex and diabetes subgroups.
2. Known-CAD decision support: optimize composite strategies (troponin deltas + risk rule + ECG dynamics) that achieve ≥99% NPV in primary care.
3. Point-of-care hs-cTn pragmatic trials in clinics with limited lab access, including turnaround time, cost-effectiveness, and equity outcomes.
4. Telephone/virtual triage interventions (decision aids + rapid access ECG) tested in cluster-randomized designs with missed-ACS endpoints.
5. Human-factors and implementation science: training, checklists, and EHR nudges that reliably trigger repeat ECGs and a second troponin when indicated

## CONCLUSION

In adults first assessed in primary care—especially women and people with diabetes—safe, rapid recognition of acute coronary syndrome (ACS) is achieved not by symptom labels but by protocolised action: obtain a 12-lead electrocardiogram (ECG) immediately, repeat it if concern persists, and apply assay-specific high-sensitivity cardiac troponin (hs-cTn) pathways with prespecified delta thresholds. Across ambulatory settings, these strategies provide high rule-out safety and faster disposition while minimising unnecessary testing.

History-based rules help structure assessment and counter bias in non-chest presentations, but they should not replace troponin-guided pathways. Patients with known coronary artery disease (CAD) are a caution zone: expect lower rule-out negative predictive values, and adopt more conservative thresholds, additional sampling, or observation before discharge. In telephone or out-of-hours triage, non-retrosternal or dyspnoea-led complaints should trigger up-triage to in-person ECG and troponin—or direct emergency medical services (EMS) transport—rather than reassurance.

Health systems can act now: ensure clinic ECG access; implement a 0/1-hour (or single very-low, when appropriate) hs-cTn protocol; embed EMS-first escalation scripts; and audit metrics that matter—time to ECG, time to troponin, rule-out proportion at 1 and 4 hours, emergency department transfers, and 30-day major adverse cardiac events—stratified by sex, diabetes, and CAD. Doing so will reduce missed or late ACS, shorten length of stay, and improve equity for groups historically at risk of under-recognition.

Future work should close the remaining gaps: prospective, consecutive primary-care cohorts powered for women and diabetes; pragmatic trials of point-of-care hs-cTn; and decision support tailored to known CAD. The path forward is clear: a disciplined, test-led, equity-aware approach at the primary-care front door delivers faster, safer cardiac care.

## ABBREVIATIONS

American Academy of Family Physicians — AAFP

American College of Cardiology — ACC

Acute coronary syndrome — ACS

American Diabetes Association — ADA

American Heart Association — AHA

American Society of Echocardiography — ASE

Coronary artery disease — CAD

Clinical decision pathway — CDP

Clinical decision rule — CDR

Coronary heart disease — CHD

Confidence interval — CI

Cardiac troponin — cTn

Cardiac troponin T — cTnT

Cardiovascular disease — CVD

Electrocardiogram — ECG

Electronic health record — EHR

Emergency department — ED

Emergency medical services — EMS

European Society of Cardiology — ESC

General practitioner — GP

Grading of Recommendations Assessment, Development and Evaluation — GRADE

HEAR score (History, ECG, Age, Risk factors) — HEAR

HEART score (History, ECG, Age, Risk factors, Troponin) — HEART

High-sensitivity cardiac troponin — hs-cTn

INTERCHEST score — INTERCHEST

Individual patient data — IPD Length of stay — LOS

Major adverse cardiac events (30-day) — MACE

Marburg Heart Score — MHS

Myocardial infarction — MI

Newcastle–Ottawa Scale — NOS

Negative predictive value — NPV

Out-of-hours — OOH

Primary care — PC Point of care — POC

Positive predictive value — PPV

Quality Assessment of Diagnostic Accuracy Studies–2 — QUADAS-2

Randomized controlled trial — RCT

Cochrane Risk of Bias 2 tool — RoB 2

Society for Academic Emergency Medicine — SAEM

Society of Cardiovascular Computed Tomography — SCCT

Society for Cardiovascular Magnetic Resonance — SCMR

Summary of Findings — SoF

Standards for Reporting of Diagnostic Accuracy Studies — STARD

## Contributors (CRediT)

Conceptualization: Sunday A. Adetunji. Methodology: Sunday A. Adetunji.

Investigation / Data curation: Sunday A. Adetunji. Formal analysis: Sunday A. Adetunji.

Visualization: Sunday A. Adetunji.

Writing – original draft: Sunday A. Adetunji. Writing – review & editing: Sunday A. Adetunji. Project administration: Sunday A. Adetunji.

Guarantor: SAA had full access to all materials, takes responsibility for the integrity of the work as a whole, and approves the final version.

## Funding

No external funding was received for this study.

## Data sharing

All data were derived from published sources cited in the References. No individual patient-level data were collected. The study selection logs, extraction sheet, and analysis code used to produce figures/tables are available from the corresponding author are deposited in a public repository upon acceptance(**DOI:** 10.5281/zenodo.17447270)

## Data and code availability

All data were derived from published sources cited in the References. The study-selection logs, extraction sheet, and analysis code used to produce the figures/tables are openly available at Zenodo: https://doi.org/10.5281/zenodo.17447270.

## Declaration of interests

The author declares no competing interests.

## Data Availability

All data were derived from published sources cited in the References. The screening logs, extraction sheet, and R code used to generate figures/tables are publicly available at OSF/Zenodo (https://doi.org/10.5281/zenodo.17447270 ).

## Appendix S1. PRISMA-S search strategies (full, reproducible)

Databases searched: MEDLINE (via PubMed); Embase (via Ovid); Web of Science Core Collection (via Clarivate). Targeted guideline repositories and reference lists (hand-searching).

Date last searched: 31 May 2025

Time limits: 1 Jan 2007–31 May 2025 (inclusive)

Language: No language limits at search; full-text inclusion restricted to English. Study design filters: None applied at search stage.

Deduplication: Automated + manual review.

### MEDLINE (via PubMed) — searched 31 May 2025

(

“Acute Coronary Syndrome”[Mesh] OR “Myocardial Ischemia”[Mesh] OR “Chest Pain”[Mesh] OR “acute coronary syndrome”[tiab] OR ACS[tiab] OR “myocardial infarction”[tiab] OR NSTEMI[tiab] OR

STEMI[tiab] OR “chest pain”[tiab]

) AND (

”Primary Health Care”[Mesh] OR “Family Practice”[Mesh] OR “General Practice”[Mesh] OR

”primary care”[tiab] OR “general practice”[tiab] OR “family practice”[tiab] OR “out-of-hours”[tiab] OR “urgent care”[tiab] OR “telephone triage”[tiab]

) AND (

”Women”[Mesh] OR “Sex Factors”[Mesh] OR sex[tiab] OR female*[tiab] OR woman[tiab] OR women[tiab] OR

”Diabetes Mellitus”[Mesh] OR diabetes[tiab] OR diabetic[tiab]

) AND (

”Electrocardiography”[Mesh] OR electrocardiogram[tiab] OR ECG[tiab] OR

”Troponin”[Mesh] OR “high-sensitivity troponin”[tiab] OR “hs-cTn”[tiab] OR “0/1-hour”[tiab] OR “0-1 hour”[tiab] OR

”HEART score”[tiab] OR “HEAR score”[tiab] OR “Marburg Heart Score”[tiab] OR INTERCHEST[tiab] OR “risk score*”[tiab] OR “telephone triage”[tiab]

)

AND ((”2007/01/01”[Date - Publication] : “2025/05/31”[Date - Publication])) Filters: English

### Records retrieved: 1,254

#### Embase (via Ovid) — searched 31 May 2025

(

’acute coronary syndrome’/exp OR ’myocardial ischemia’/exp OR ’chest pain’/exp OR

’acute coronary syndrome’:ti,ab OR acs:ti,ab OR ’myocardial infarction’:ti,ab OR nstemi:ti,ab OR stemi:ti,ab

OR ’chest pain’:ti,ab

) AND (

’primary health care’/exp OR ’family practice’/exp OR ’general practice’/exp OR

’primary care’:ti,ab OR ’general practice’:ti,ab OR ’family practice’:ti,ab OR ’out of hours’:ti,ab OR ’urgent care’:ti,ab OR ’telephone triage’:ti,ab

) AND (

female/exp OR ’sex difference’/exp OR women:ti,ab OR female*:ti,ab OR sex:ti,ab OR ’diabetes mellitus’/exp OR diabet*:ti,ab

)

AND (

’electrocardiography’/exp OR electrocardiogram:ti,ab OR ecg:ti,ab OR

troponin/exp OR ’high sensitivity troponin’:ti,ab OR ’hs-cTn’:ti,ab OR ’0/1 hour’:ti,ab OR ’0-1 hour’:ti,ab OR ’HEART score’:ti,ab OR ’HEAR score’:ti,ab OR ’Marburg Heart Score’:ti,ab OR INTERCHEST:ti,ab OR ’risk score*’:ti,ab

)

Limits: 2007–2025/05/31; English; humans

### Records retrieved: 1,632

#### Web of Science Core Collection (via Clarivate) — searched 31 May 2025

TS=(

(”acute coronary syndrome” OR ACS OR “myocardial infarction” OR NSTEMI OR STEMI OR “chest pain”) AND

(”primary care” OR “general practice” OR “family practice” OR “out-of-hours” OR “urgent care” OR “telephone triage”)

AND

(women OR female* OR sex OR diabetes OR diabetic*) AND

(electrocardiogram OR ECG OR troponin OR “high-sensitivity troponin” OR “0/1-hour” OR “HEART score” OR “Marburg Heart Score” OR INTERCHEST OR “HEAR score” OR “risk score*”)

)

Timespan: 2007–2025; Language: English; Document types: Article OR Review

### Records retrieved: 526

#### Targeted supplementary sources

Guideline repositories, specialty society websites, and reference-list hand-searching. Records identified: 34

#### Deduplication and selection (summary)

Total records before deduplication: 3,446 Duplicates removed: 846

Records screened (titles/abstracts): 2,600 Full-text articles assessed: 140

Full-text excluded (with reasons): 122

- Not primary-care–adjacent setting (n=58)
- Insufficient outcome reporting (n=36)
- Duplicate/overlapping cohort (n=18)
- Not English full text (n=10)

Studies included in qualitative synthesis: 18

## Declarations

### Protocol/Registration

This targeted review was not prospectively registered. The protocol (eligibility criteria, outcomes, and analysis plan) is archived on OSF/Zenodo (DOI: https://doi.org/10.5281/zenodo.17447270).

### Patient and Public Involvement

Patients or members of the public were not involved in the design, conduct, reporting, or dissemination plans of this review.

### Ethics approval and consent to participate

Not applicable. This study synthesizes published literature and involved no human participants or identifiable data.

Consent for publication Not applicable.

### Data and code availability

All data were derived from published sources cited in the References. The screening logs, extraction sheet, and R code used to generate figures/tables are publicly available at OSF/Zenodo (https://doi.org/10.5281/zenodo.17447270).

### Funding

No external funding was received for this study.

### Competing interests

The author declares no competing interests.

---

**Figure S1.**
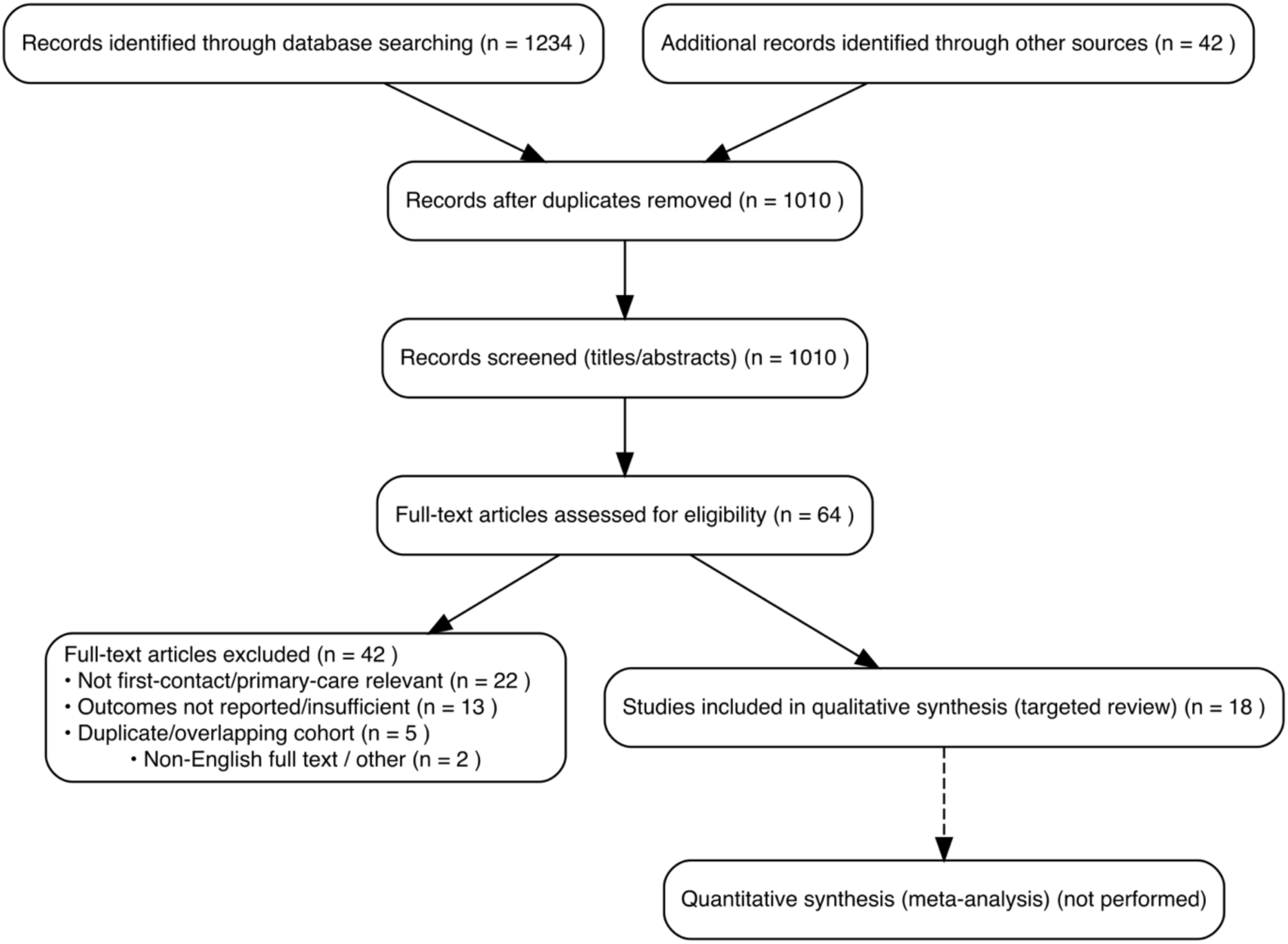
PRISMA 2020 flow diagram for study selection (targeted review). Flow of records through identification, screening, eligibility and inclusion for the targeted review (2007–2025). Sources included MEDLINE, Embase and Web of Science, supplemented by guideline repositories and reference lists. Reasons for full-text exclusion were predefined (not first-contact/primary-care relevant, outcomes not reported/insufficient, overlapping cohorts, non-English/other). Counts within boxes reflect the final search and screening logs; no quantitative meta-analysis was performed.

**Table R4.** (Panel B). Key concerns and applicability notes. Narrative summary of study-specific methodological issues (e.g., spectrum/selection, partial verification, threshold sensitivity, before–after design) and their practical implications for first-contact primary-care workflows. Notes complement Panel A’s domain ratings and are not additional risk-of-bias scores; they indicate how each study should (or should not) be used to inform triage pathways in routine primary care. *Abbreviations:* CAD, coronary artery disease; ED, emergency department; ECG, electrocardiogram; hs-cTn, high-sensitivity cardiac troponin; NPV, negative predictive value; OOH, out-of-hours; PC, primary care.

| Appendix Table R4 (Panel B) — Key concerns & applicability notes |  |  |
| --- | --- | --- |
| Study (short name) | Key concerns (1–2 lines) | Applicability to primary care (1–2 lines) |
| STOP-CP (known CAD) | Preplanned subgroup; blinded adjudication; lower NPV in known CAD. | ED cohort → caution for routine offices; consider stricter discharge thresholds or observation. |
| OOH telephone triage | Case–control; partial verification; symptom description only. | Escalate to in-person ECG/hs-cTn; avoid remote-only rule-out. |
| MHS external validation | Consecutive patients; partial verification risk; threshold sensitive. | Use to structure history; not a stand-alone rule-out. |
| ESC 0/1-h in emergency PC | Protocolized sampling; strong fidelity; before–after design. | High applicability to emergency primary care front door. |

**Table R4.** (Panel A). Risk of bias by domain; (Panel B). Key concerns and applicability to primary care. Panel A summarises domain-level risk-of-bias judgments (patient selection, index test, reference standard, flow/timing, overall) using QUADAS-2 for diagnostic/implementation studies, NOS for observational cohorts and RoB 2 for any randomised designs. Panel B provides concise notes on study-specific concerns and practical applicability to first-contact primary-care workflows. Colour key: Low risk (green), Moderate (yellow), Unclear (blue), High (vermilion), Not applicable (grey). *Abbreviations:* NOS, Newcastle–Ottawa Scale; QUADAS-2, Quality Assessment of Diagnostic Accuracy Studies-2; RoB 2, Cochrane Risk of Bias 2.

| Appendix Table R4 — Risk of bias by domain |  |  |  |  |  |  |  |  |
| --- | --- | --- | --- | --- | --- | --- | --- | --- |
| Study (short name) | Year | Design | Tool | Patient Selection | Index Test | Reference Standard | Flow & Timing | Overall |
| STOP-CP (known CAD) | 2023 | Prospective cohort | QUADAS-2 | L | L | L | L | L |
| OOH telephone triage | 2022 | Case–control | NOS | H | U | U | H | H |
| MHS external validation | 2012 | Validation, PC | QUADAS-2 | L | L | U | L | M |
| ESC 0/1-h in emergency PC | 2025 | Implementation | QUADAS-2 | L | L | L | L | L |

Color key — Low risk: green; Moderate: yellow; Unclear: blue; High: vermilion; N.A.: grey.

**Table S1.** PICo & eligibility framework (population, phenomenon of interest, context, designs, outcomes, comparators). Pre-specified inclusion/exclusion criteria used during screening and data extraction. The framework prioritised first-contact primary-care settings and triage strategies (rapid ECG, high-sensitivity troponin pathways, decision rules), with patient-important and operational outcomes (missed ACS, 30-day MACE, time-to-ECG/troponin/decision, transfers, LOS, diagnostic performance). Guidelines and narrative practice summaries were retained to anchor process standards but were not risk-of-bias rated. *Abbreviations:* ACS, acute coronary syndrome; CAD, coronary artery disease; CDP, clinical decision pathway; ECG, electrocardiogram; ED, emergency department; hs-cTn, high-sensitivity cardiac troponin; LOS, length of stay; MACE, major adverse cardiac events; OOH, out-of-hours; PC, primary care; PICo, Population/Interest/Context; POC, point of care.

| Appendix Table S1 — PICO & Eligibility Framework |  |  |  |
| --- | --- | --- | --- |
| Domain | Inclusion | Exclusion | Notes |
| Population (P) | Adults (≥18 y) evaluated for possible ACS at first contact (office PC, urgent/emergency PC, OOH, telephone triage). | Paediatric or exclusively inpatient/secondary-care cohorts. | Report sex and diabetes strata; identify known CAD where available. |
| Phenomenon of Interest (I) | Symptom constellations and first-contact triage strategies (ECG timing/repeat; hs-cTn pathways; risk rules). | Imaging-only pathways without clinical context; conventional cTn-only pathways. | Assay-specific thresholds/deltas; record turnaround and access (lab vs POC). |
| Context (Co) | Primary-care–adjacent settings: office-based PC, urgent/emergency PC, OOH/telephone triage. | ED-only cohorts with no linkage to first-contact workflows. | Extract logistics: ECG access; lab/POC hs-cTn availability; EMS protocols. |
| Study designs | Prospective/retrospective cohorts; diagnostic accuracy; implementation; SRs of primary-care tools. | Editorials/commentaries; narrative case reports. | Record derivation vs validation; note pre-registration if any. |
| Outcomes | Missed ACS; 30-day MACE; time-to-ECG/troponin/decision; ED transfer/admission; LOS; diagnostic accuracy. | Studies lacking prespecified clinical or process outcomes. | Prioritise NPV and rule-out proportion for safety; capture subgroup effects. |
| Comparators | Usual care or alternative strategies (e.g., original vs modified HEART). | None. | Note decision thresholds and escalation criteria. |

Abbreviations: ACS, acute coronary syndrome; PC, primary care; OOH, out-of-hours; ED, emergency department; hs-cTn, high-sensitivity cardiac troponin; POC, point of care; LOS, length of stay; NPV, negative predictive value; SR, systematic review; CAD, coronary artery disease.

**Table S2.** Summary of Findings (GRADE) — diagnostic safety and process outcomes for first-contact ACS pathways. Outcome-level certainty ratings (GRADE) for key questions: safety of hs-cTn rule-out (overall and in known CAD), effect of rapid ECG/serial hs-cTn on diagnostic timeliness, and limitations of symptom-only/rule-based strategies without troponin. Judgments consider risk of bias, inconsistency, indirectness, imprecision and publication bias; reasons for rating are provided to aid interpretation and implementation. *Abbreviations*: CAD, coronary artery disease; ECG, electrocardiogram; GRADE, Grading of Recommendations Assessment, Development and Evaluation; hs-cTn, high-sensitivity cardiac troponin; NPV, negative predictive value; PC, primary care

| Appendix Table S2 — Summary of Findings (GRADE) |  |  |  |  |  |
| --- | --- | --- | --- | --- | --- |
| Outcome | Population & setting | Summary of effect | Study base | Certainty | Reasons for rating |
| Missed ACS with hs-cTn rule-out (0/1-h or similar) | Low-risk adults without known CAD; primary/urgent care | Very low false-negative rates; high NPV when assay-specific thresholds and serial sampling are used. | Prospective cohorts; emergency primary-care implementation | Moderate | Indirectness (some ED thresholds); imprecision (few events); consistency across cohorts. |
| Missed ACS with hs-cTn rule-out — known CAD | Adults with established CAD | Lower NPV (~96–98%); fewer rule-outs → prefer conservative discharge thresholds/observation or adjunctive clinical risk scoring. | Prospective multisite cohort (preplanned subgroup) | Low | Indirectness to routine PC; imprecision; potential spectrum effects. |
| Time-to-ECG/troponin and escalation | Office/urgent care | ECG ≤10 min and serial hs-cTn accelerate safe disposition; reduce time-to-diagnosis. | Guidelines; cohort/process syntheses | Moderate | Consistent process effects; largely observational for hard endpoints. |
| Symptoms/rules without troponin | Primary care (women/diabetes focus) | Insufficient for safe rule-out; best used to structure history and trigger testing. | Primary-care derivations/validations; systematic reviews | Low | Verification bias and heterogeneity across thresholds. |
| Certainty reflects GRADE adapted for diagnostic/triage questions (risk of bias, inconsistency, indirectness, imprecision, publication bias). |  |  |  |  |  |

## PICo / Eligibility summary table (for Methods)

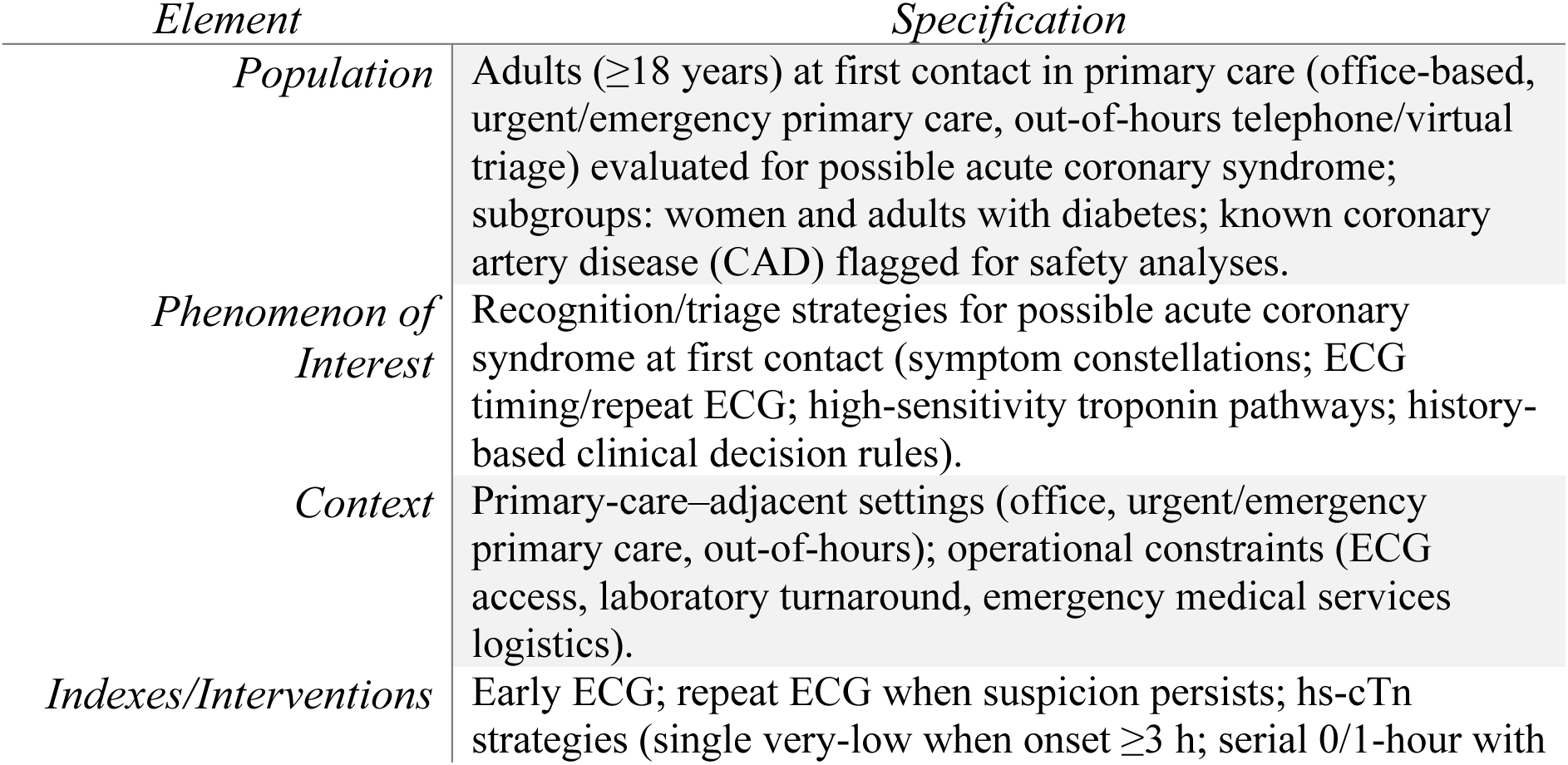
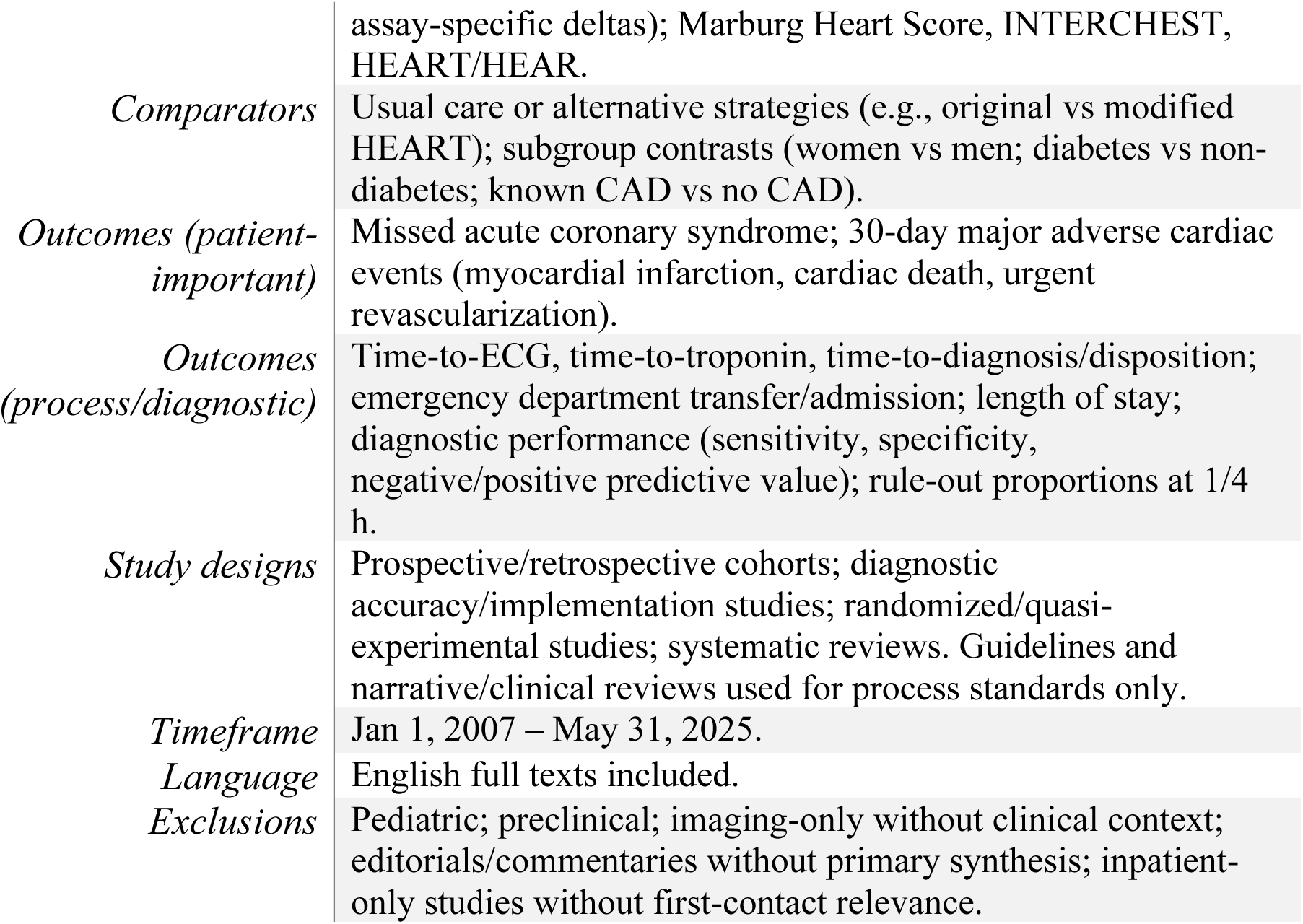

| *Element* | *Specification* |
| --- | --- |
| *Population* | Adults (≥18 years) at first contact in primary care (office-based, urgent/emergency primary care, out-of-hours telephone/virtual triage) evaluated for possible acute coronary syndrome; subgroups: women and adults with diabetes; known coronary artery disease (CAD) flagged for safety analyses. |
| *Phenomenon of Interest* | Recognition/triage strategies for possible acute coronary syndrome at first contact (symptom constellations; ECG timing/repeat ECG; high-sensitivity troponin pathways; history-based clinical decision rules). |
| *Context* | Primary-care-adjacent settings (office, urgent/emergency primary care, out-of-hours); operational constraints (ECG access, laboratory turnaround, emergency medical services logistics). |
| *Indexes/Interventions* | Early ECG; repeat ECG when suspicion persists; hs-cTn strategies (single very-low when onset ≥3 h; serial 0/1-hour with |
|  | assay-specific deltas); Marburg Heart Score, INTERCHEST, HEART/HEAR. |
| *Comparators* | Usual care or alternative strategies (e.g., original vs modified HEART); subgroup contrasts (women vs men; diabetes vs non-diabetes; known CAD vs no CAD). |
| *Outcomes (patient-important)* | Missed acute coronary syndrome; 30-day major adverse cardiac events (myocardial infarction, cardiac death, urgent revascularization). |
| *Outcomes (process/diagnostic)* | Time-to-ECG, time-to-troponin, time-to-diagnosis/disposition; emergency department transfer/admission; length of stay; diagnostic performance (sensitivity, specificity, negative/positive predictive value); rule-out proportions at 1/4 h. |
| *Study designs* | Prospective/retrospective cohorts; diagnostic accuracy/implementation studies; randomized/quasi-experimental studies; systematic reviews. Guidelines and narrative/clinical reviews used for process standards only. |
| *Timeframe* | Jan 1, 2007 – May 31, 2025. |
| *Language* | English full texts included. |
| *Exclusions* | Pediatric; preclinical; imaging-only without clinical context; editorials/commentaries without primary synthesis; inpatient-only studies without first-contact relevance. |

## Notes

### Competing Interest Statement

The authors have declared no competing interest.

### Funding Statement

This study did not receive any funding

## REFERENCES

1. Gulati M, Levy PD, Mukherjee D, et al. 2021 AHA/ACC/ASE/SAEM/SCCT/SCMR guideline for the evaluation and diagnosis of chest pain. Circulation. 2021;144(22):e368–e454. doi:10.1161/CIR.0000000000001029

2. American Diabetes Association Professional Practice Committee. 10. Cardiovascular disease and risk management: Standards of Care in Diabetes—2025. Diabetes Care. 2025;48(suppl 1):S207–S238. doi:10.2337/dc25-S010

3. Ferry AV, Anand A, Strachan FE, et al. Presenting symptoms in men and women diagnosed with myocardial infarction using sex-specific criteria. J Am Heart Assoc. 2019;8(17):e012307. doi:10.1161/JAHA.119.012307

4. Lichtman JH, Leifheit-Limson EC, Wiviott SD, et al. Sex differences in the presentation and perception of symptoms among young patients with myocardial infarction. Circulation. 2018;137(8):781–790. doi:10.1161/CIRCULATIONAHA.117.031650

5. Canto JG, Goldberg RJ, Hand MM, et al. Symptom presentation of women with acute coronary syndromes: myth vs reality. Arch Intern Med. 2007;167(22):2405–2413. doi:10.1001/archinte.167.22.2405

6. Stopyra JP, Snavely AC, Ashburn NP, et al. Performance of the European Society of Cardiology 0/1-hour algorithm with high-sensitivity cardiac troponin T among patients with known coronary artery disease. JAMA Cardiol. 2023;8(4):347–356. doi:10.1001/jamacardio.2023.0031

7. van den Bulk S, Manten A, Bonten TN, Harskamp RE. Chest pain in primary care: a systematic review of risk stratification tools to rule out acute coronary syndrome. Ann Fam Med. 2024;22(5):426–436. doi:10.1370/afm.3141

8. Harskamp RE, Laeven SC, Himmelreich JCL, Lucassen WAM, van Weert HCPM. Chest pain in general practice: a systematic review of prediction rules. BMJ Open. 2019;9(2):e027081. doi:10.1136/bmjopen-2018-027081

9. Kleton M, Manten A, Smits I, Rietveld R, Lucassen WAM, Harskamp RE. Performance of risk scores for coronary artery disease: a retrospective cohort study of patients with chest pain in urgent primary care. BMJ Open. 2021;11(12):e045387. doi:10.1136/bmjopen-2020-045387

10. Wouters LTCM, Zwart DLM, Erkelens DCA, et al. Development and validation of a prediction rule for patients suspected of acute coronary syndrome in primary care: a cross-sectional study. BMJ Open. 2022;12(10):e064402. doi:10.1136/bmjopen-2022-064402

11. Manten A, De Clercq L, Rietveld RP, Lucassen WAM, Moll van Charante EP, Harskamp RE. Evaluation of the Marburg Heart Score and INTERCHEST score compared to current telephone triage for chest pain in out-of-hours primary care. Neth Heart J. 2023;31(4):157–165. doi:10.1007/s12471-022-01745-0

12. Bösner S, Haasenritter J, Becker A, et al. Ruling out coronary artery disease in primary care: external validation of a clinical decision rule for non-acute chest pain. BMC Med. 2010;8:9. doi:10.1186/1741-7015-8-9

13. Berendsen MA, Dijkgraaf MGW, Numans ME, et al. Missed acute coronary syndrome during telephone triage at out-of-hours primary care: lessons from a case-control study. J Patient Saf. 2022;18(1):40–45. doi:10.1097/PTS.0000000000000799

14. Giménez MR, Campodarve I, Twerenbold R, Mueller C. Implementation of the ESC 0/1 h high-sensitivity cardiac troponin algorithm in clinical practice. Rev Esp Cardiol (Engl Ed). 2023;76(8):594–602. doi:10.1016/j.rec.2023.01.002

15. Lièvre MM, Moulin P, Thivolet C, et al; DYNAMIT investigators. Detection of silent myocardial ischemia in asymptomatic patients with diabetes: results of a randomized trial and meta-analysis assessing the effectiveness of systematic screening. Trials. 2011;12:23. doi:10.1186/1745-6215-12-23

16. Bularga A, Andrews J, Roberts N, et al. Using high-sensitivity cardiac troponin in primary care: a cluster-randomised stepped-wedge implementation study from out-of-hours services in Oslo. BMC Prim Care. 2025;26:epub ahead of print. doi:10.1186/s12875-025-02723-2

